# Patient experiences of UK primary care online triage and consultation platforms during COVID-19: A systematic review

**DOI:** 10.1101/2024.01.09.24301062

**Authors:** Christopher Roberts, Jomin George, Judy Jenkins

## Abstract

**Background:** Online triage and consultation platforms are being increasingly used by healthcare providers in the UK for patient/clinician interactions. COVID-19 accelerated the shift towards the use of these platforms to maintain healthcare provision and reduce transmission. Strict directives were introduced by the UK Government to avoid in-person contact wherever possible in March 2020.

**Aim:** To examine patients’ experiences of online triage and consultation in UK primary care during the COVID-19 pandemic and offer considerations for their continued use.

**Design:** This study follows the PRISMA framework and includes qualitative studies conducted in UK primary care based on the experiences of patient users of any such online platform during the period of March 2020 to April 2023. Studies were included using the PICO format. Three literature databases were searched for relevant studies: PubMed, Science Direct and EMBASE. CASP is used to assess data quality.

**Results:** 540 studies were reviewed and reduced to 12 studies that met the inclusion criteria. Study characteristics were identified as: year of study, study population, disease types/conditions, patient response themes and the study’s data capture method. A thematic inductive approach identifies three overarching themes (Accessibility, Care delivery, System functionality) and 10 sub-themes (Affordability, IT literacy, Communication, Convenience, Care quality, Patient safety/privacy, Usability, Continuity of care, Inequality and Media influence).

**Conclusion:** This review highlights aspects of patient satisfaction and benefit but also those most concerning for patients. This study reviews the rapid, compulsory adoption of these systems during COVID-19 with implications for their future implementation beyond the pandemic.

## Introduction

The use of online triage and consultation platforms is increasingly used amongst healthcare providers in the UK.^1^ These platforms are provided by private sector vendors, from an approved National Health Service (NHS) supplier list, and they allow patients to remotely communicate, synchronously or asynchronously, with a healthcare practitioner.^2^ Key features of the online platforms can include message exchange, appointment booking, symptom checking, photo sharing, signposting to other services and direct video consultation^3,4^.

The use of these online tools has been driven by increasing workload pressures facing the NHS, increasing patient acceptance of and accessibility to technology, and a downward trend in patient satisfaction^5,6^.

During the COVID-19 pandemic, the shift towards the use of these platforms was accelerated due to viral transmission mitigation measures introduced by the UK Government and guidance from healthcare authorities to implement more of these remote systems to maintain levels of healthcare provision^7,8^. By April 2020, 90% of all consultations in primary care were delivered remotely^9^.

## Results

The literature search yielded a total of 540 results. 12 studies met the inclusion criteria using the PRISMA checklist framework ^20^.

### Study characteristics

UK-based studies were limited. The studies spanned a pandemic timeframe between January 2020 and April 2023. Populations represented in the studies were broadly inclusive and predominantly undertaken within the primary care landscape of England. Four of the twelve studies examined patient experiences within specific disease or condition types. 10 themes emerged. In all but one of the studies patient experience data was captured either by survey, interview or focus group. In the exception, data was gathered from social media comments posted on the platform Twitter (now known as X). Study characteristics are captured in Table 3.

### Data quality

Seven out of eight studies of the academic literature sources used in this review were sourced from journals with a high impact factor, ranging from 2.908 to 7.077. The four grey literature sources used are credible^21,22^. A quality appraisal of the selected literature was undertaken using the CASP framework (Table 4). Risk of bias was identified in five of the 12 selected studies.

### Data analysis

This review adopts a data-led, inductive thematic analysis framework to derive themes and codes reflective of the data represented in the studies^35,36^. The data were categorised into three overarching themes with 10 sub-themes under each. The frequency of each thematically coded data point is shown in the QDA Miner extract (figure 2).

**Figure 1.**
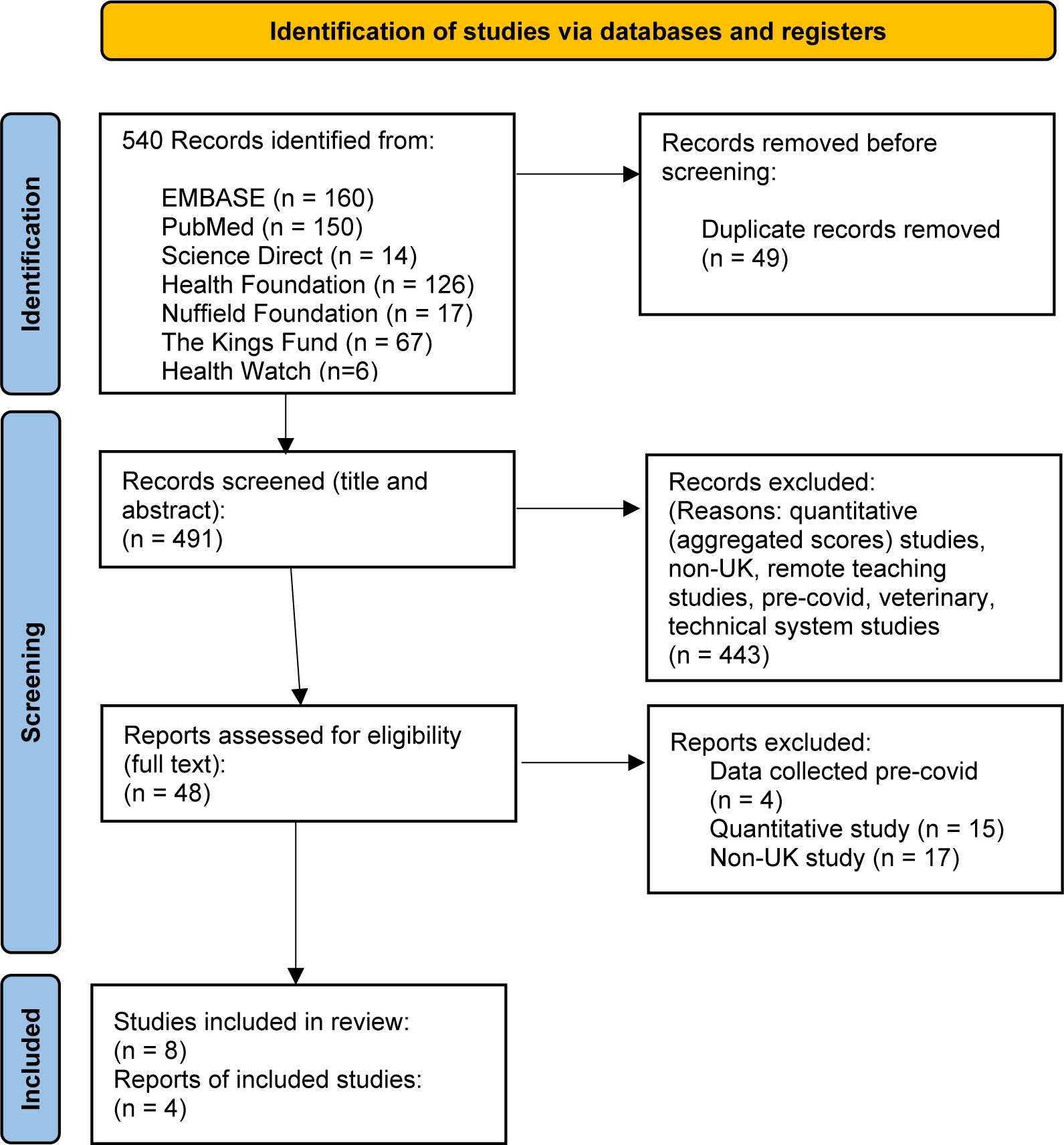
PRISMA Flow Diagram of study selection.

**Figure 2.**
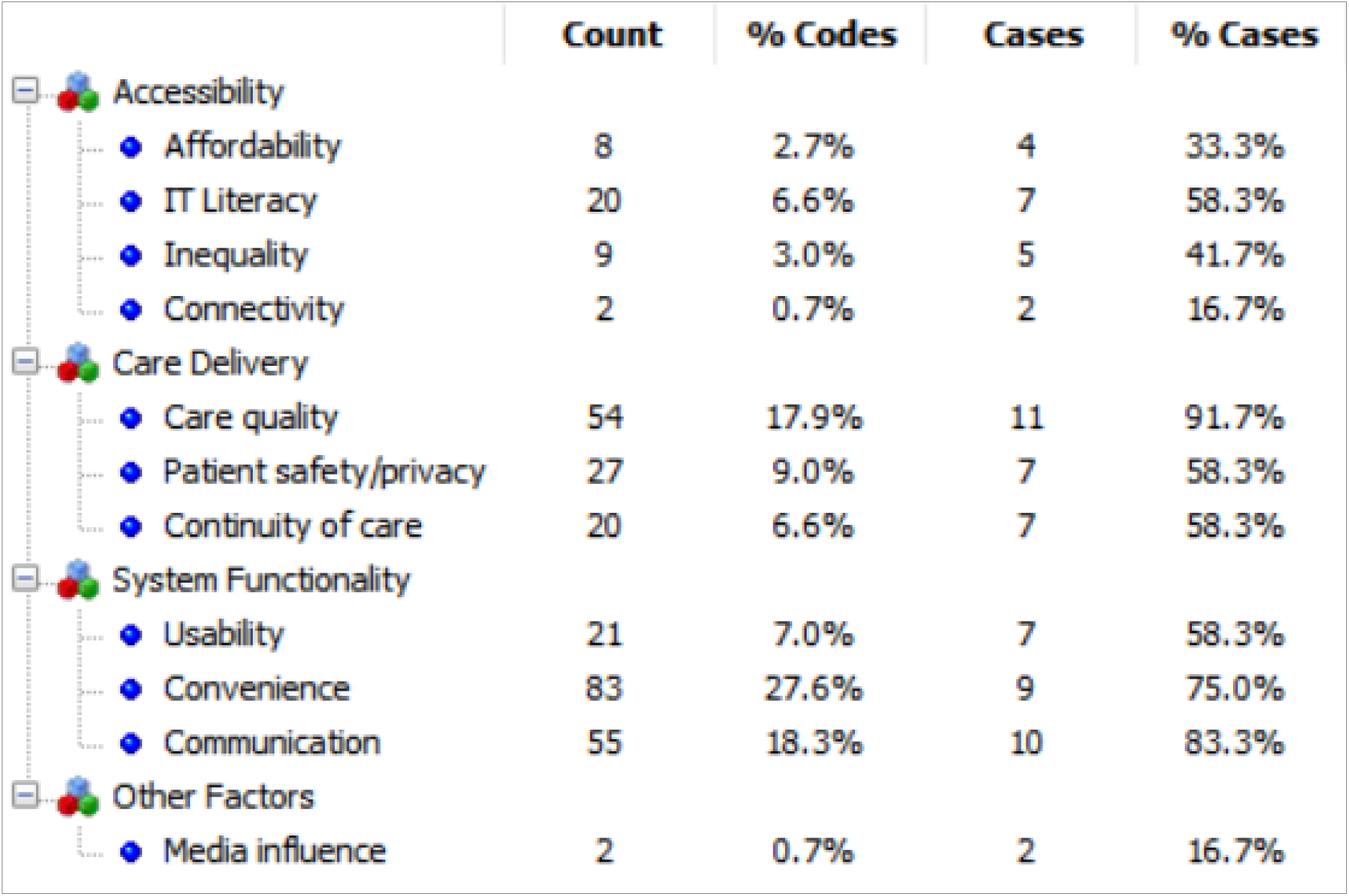
Number of data points by theme using QDA Miner.

### Main themes

#### Accessibility - Affordability

Eight data points around the theme of Affordability were revealed. This theme was most prominent in Verity^34^ who examined the remote primary care experiences of minority and high-need groups (Gypsy and Travellers, sex workers, vulnerable migrants, and those experiencing homelessness). Within the 21 data points, respondents reported financial barriers to access due to having non-contract, ‘top-up’ mobile phones with limited credit and limited data allowance to transfer multi-media files, such as photographs, requested by GPs to adequately provide care^34^. Affordability crossed over with the theme of IT Literacy, with one participant stating that even with access to technology they did not possess the skills to effectively use it.

In Howells^25^ (experiences of homeless people), similar themes emerged from two participants. One respondent reported ‘on hold’ phone delays at their GP would use up available call credit and another that a shared computer managed by a homeless hostel was unavailable due to Covid restrictions^25^. Similarly, in HealthWatch^27^ one participant expressed cost constraints over call waiting times with their GP after being unable to use the online booking system.

#### Accessibility - IT Literacy

The theme of IT Literacy featured most prominently in HealthWatch^27^ and Verity^34^, the former a study consisting of a high proportion of over 60s (63%) and the latter consisting of marginalised groups. Every response under this theme in HealthWatch^27^ stated a reliance on someone else, predominantly a family member, to use the online systems with one participant suggesting the implementation of a volunteer-led programme of support. The need for support for elderly patients unfamiliar with technology was echoed by one participant in Turner^33^, as well as a social media comment captured in Ainley^23^. In Verity^34^ similar support was sought from friends, interpreters or support workers. Duncan^24^ reported a patient using online system instructions from another GP practice to access their own GPs system effectively. A HealthWatch^27^ participant highlighted IT illiteracy, exacerbated by a language barrier, as the reason they were unable to navigate their GPs online system.

#### Accessibility – Inequality

All four participants coded under Inequality in Verity^34^ expressed frustration that online systems marginalised them further for reason of a lack of formal identification to use the health service, low written literacy levels and a perceived discriminatory reluctance for physical health examination. Inequalities over access to care were also raised by breastfeeding women, mental health patients^31^ the elderly^23,32^, dementia patients and blind or deaf patients without interpretation assistance.^27^

#### Care Delivery - Care Quality

The theme of care quality was the third highest reported in the literature and was evident in 11 out the 12 studies included. This theme featured heavily (23 out of 54 responses) in Oxleas^32^ patient survey of health service users in the Southeast London region during the first few months of the pandemic. Participants broadly felt the remote delivery of care matched that of in-person care quality. This perception was aided by the availability of a video consultation option with several respondents positively reporting that this enabled them to effectively communicate symptoms and pain, removed stress and anxiety of in-person attendance and delivered satisfactory treatment and outcomes^32^. This was balanced by some negative perceptions that included: an aggravation of mental health conditions, a lack of emotional connection and an over-reliance on outdated patient records which, due to a lack of physical examination, were perceived as not reflecting patients’ current condition.

Elsewhere in the literature some patients commented positively on GPs transition to the remote model^27^, whilst negative care quality comments related to impersonal communication^28^, lack of empathy^25^ and no access to physical examination^32,34^. Thematic cross-over was revealed between Care Quality and Continuity of Care whereby a lack of the latter negatively affected the former in Ladds^30^. Notably, one respondent in Ladds^30^ recognised a pattern in the online system’s logic processing to ensure the query was met with a direct GP response as opposed to being redirected to other support services. The patient felt it necessary to knowingly enter inaccurate health information which subsequently led to concern over the care that was received.

#### Care Delivery - Patient Safety/Privacy

10 participants across the studies who gave positive responses related it to their reluctance to attend in-person appointments, which were exacerbated by COVID-19, due to having mobility constraints, chronic tiredness, frequent panic attacks or other multi-morbidities.

Three respondents reacted positively to patient safety citing health benefits of not being potentially exposed to the COVID-19. Three patients were concerned about their privacy when using online consultation platforms; two relating to hesitancy using the platform at home for fear of revealing sensitive health information and another with concerns over who would have access to their health information once submitted to the online system.

#### Care Delivery - Continuity of Care

Seven patients experienced frustration or dismay at having to explain, often complex, health history as the online system did not allow patients to seek care from a familiar practitioner, with one patient in Health Foundation^31^ who perceived patients were seen as “faceless creatures”. Conversely, five patients relayed positive continuity of care sentiments where online systems allowed patients to consult with a GP familiar to them and their health history. Notably, one respondent in Health Foundation^31^ reported using an online system with a published rota of practitioner availability but found it was not always accurate.

#### System Functionality – Usability

Seven of the 12 included studies surfaced comments relating to the usability and functionality of the online system. The frequency of positive (6) and negative (6) patient views were balanced in the studies. Elements of frustration included a lack of system notifications, unfamiliar software within the bespoke platforms, restrictions on certain types of devices, the use of jargon, and disparate points of access requiring multiple login credentials. Causation of positive Usability views were unclear, for example, one participant commented, ‘it’s simple’^34^. Other positive sentiments related to the ability to upload attachments and the availability of support resources such as YouTube links.^27^

One contentious system feature was the facility to enter triage information by text before video consultation. Three patients commented negatively that the triage questioning was unnecessarily ‘very lengthy’^27^, ‘quite laborious’^32^ and ‘death by a thousand questions’^28^. Contrary to these statements, two patients felt the process meant they were more ‘prepared’ and had enabled the patient to ‘collect [their] thoughts’ prior to consultation.^27^ In relation to information gathering prior to consultation, two patients perceived the process as redundant if practitioners already hold their medical records.^27^

#### System Functionality – Convenience

This was the most prominent factor throughout all of the selected literature with 83 respondents alluding to it and featured in nine out the 12 studies. Comments were largely positive citing reasons of ease, speed, efficiency, and affording patients the ability to fit appointments around family and work commitments. The largest volume of comments around time savings related to a reduction in travel times and waiting room attendances. Only one patient conveyed a negative response to ‘convenience’ which related to at-work restrictions on accessing an online device^28^.

#### System Functionality – Communication

Communication was the second most prominent factor and featured in 10 of the 12 studies. Comments were overwhelmingly negative in tone with patients referencing communication deficiencies in the transition to the triage and consultation processes during COVID-19, a lack of advanced notice before appointments, no indication of when preferred practitioner was available, inability to get through by phone when experiencing system problems, and predominantly, a lack of physical communication cues when compared to face-to-face consultations. Only three positive comments were retrieved to suggest that the online platform was equitable to face-to-face contact with their GP. A notable thematic cross-over was observed between Communication and Patient Safety/Privacy and Care Quality, whereby the former impacted negatively on the latter two factors.

#### Media Influence

One participant in Ainley^23^ delayed contact with primary care due to a media-influenced perception of an over-stretched health service and another in HealthWatch^28^ thought face-to-face GP contact should resume in-line with other businesses following a media-reported relaxation of ‘lockdown’ rules.

## Discussion

### Summary

Examination of the 12 studies included in this review presented no clear evidence that patients’ experiences were generally positive or negative. This review’s thematic analysis, however, does indicate elements of satisfaction or frustration through an inductive thematic investigation that provides useful considerations for future online platform development.

The largest number of positive patient responses in the literature fell under theme of ‘Convenience’. The majority of these responses were seen in Oxleas^32^, a study conducted during the early phase of the pandemic. Due to the swift adoption of online systems, at scale, this intervention may have been seen as a novel approach to primary healthcare delivery ^9^, contributing to positive sentiments around ‘convenience’. The largest study included in this review, Oxleas^32^ captured responses from, principally, the 35-54 age demographic, many of whom welcomed the implementation of online GP access, citing reasons of convenience due to work and family commitments. Whereas in Verity^34^ and Turner^33^, studies which recruited older patients, the theme of convenience failed to feature. These studies were also conducted later during the pandemic, possibly once the novelty of online GP access had somewhat waned.

This review has also highlighted regional variation within the same themes. In HealthWatch^28^, patients from Northern and Southern counties of England reported varying care quality perceptions of their regional online systems. Different responses were also evident depending on socio-demographic factors. Two of the studies^25,34^ captured responses from marginalised groups where themes of affordability and inequality featured prominently in contrast to the other included studies.

### Strengths

- This is the first time online triage and consultation systems have been systematically reviewed based on qualitative, experiential data from patients during COVID-19.
- The literature sourced for this review contained data from multiple modalities of data capture (surveys, focus groups, interviews).
- Includes data on high-need health groups.
- The selected literature provided evidence from across the significant periods of the COVID-19 pandemic timeframe (March 2020 to May 2023).

### limitations

- Small volume of patient experience data available perhaps indicative of the ‘all-hands-on-deck’ approach to swiftly containing Covid-19 or limited access to patients for research purposes^42^.
- Some socio-demographic and specific health conditions not accounted for in the available literature.
- Limited evidence from the UK’s devolved nations which could overlook variances as health care delivery is devolved, however, evidence suggests a similar approach was adopted across nations^37^.

### Recommendations

These interconnected themes should be considered for the longevity and successful development of online platforms. This review provides crucial evidence towards reversing a stark decline in levels of patient satisfaction with healthcare services^38^. Public trust is important for healthcare and public bodies because it can improve population health through increased patient engagement with consultations, treatments and therapies thereby reducing costs and improving efficiency^39^. An absence of public trust can also contribute towards vaccine hesitancy, making it difficult for public authorities to manage public health crises, such as COVID-19, in the future^40^.

The broad range of themes presented suggests a wholistic approach to online care delivery is needed, one that considers system selection and adoption, staff training for preparedness and adequate communication with patients about process changes. More data pertaining to patient experience to facilitate further research would be advantageous.

## Method

### Study design

This systematic review follows the PRISMA study design framework which establishes a robust approach to scientific literature review^10,11,12^.

### Inclusion and exclusion criteria

This review includes qualitative studies conducted in the UK primary care landscape that have generated data based on the experiences of patient users of any online triage and consultation platform during the COVID-19 pandemic period of March 2020 to April 2023. Literature was assessed for inclusivity using the PICO framework (Table 1).

**Table 1.** PICO Framework of inclusion and exclusion criteria.

| PICOs | Inclusion Criteria | Exclusion Criteria |
| --- | --- | --- |
| Population | UK primary care patients | GPs, nurses, receptionists or other clinical staff |
| Intervention | Direct or indirect experience of using any online triage or consultation tool during the COVID-19 pandemic | Patients with no knowledge of using online triage or consultation tools during COVID-19 and patients with solely pre-COVID-19 experiences. |
| Comparison | None - No comparison is made with pre-COVID-19 use of these tools | N/A |
| Outcome | First-hand experiences, positive or negative, of using the intervention | Aggregated, quantitative scores of qualitative patient responses. |

### Information sources and search strategy

Three databases were searched: PubMed, Science Direct and EMBASE. Sources of grey literature included The Nuffield Trust, The Kings Fund, The Health Foundation and Health Watch. Searches were conducted between March and April 2023. The search term ‘Covid’ was used as a proxy for restricting by date. A combination of Boolean operators and specific phrase search terms were used for each database (Table 2).

**Table 2.** Search terms for each literature source database.

| **PubMed and EMBASE literature search terms:** |
| --- |
| ("online triage"[Title/Abstract] OR "online consultation"[Title/Abstract] OR "telehealth"[Title/Abstract] OR "remote consultation"[Title/Abstract]) AND ("covid"[Title/Abstract] OR "coronavirus"[Title/Abstract] OR "SARS-COV-2"[Title/Abstract]) AND ("primary care"[Title/Abstract] OR "general practice"[Title/Abstract]) AND ("experience"[Title/Abstract] OR "experiences"[Title/Abstract] OR "views"[Title/Abstract] OR "perceptions"[Title/Abstract]) |
| **Science Direct literature search terms:** |
| ("online triage" OR "online consultation" OR "telehealth" OR "remote consultation") ("covid" OR "Coronavirus" OR "SARS-COV-2") ("primary care" OR "general practice") ("experience" OR "views" OR "perspective") |
| **Grey literature search terms:** |
| 'primary care covid experience' |

**Table 3.**
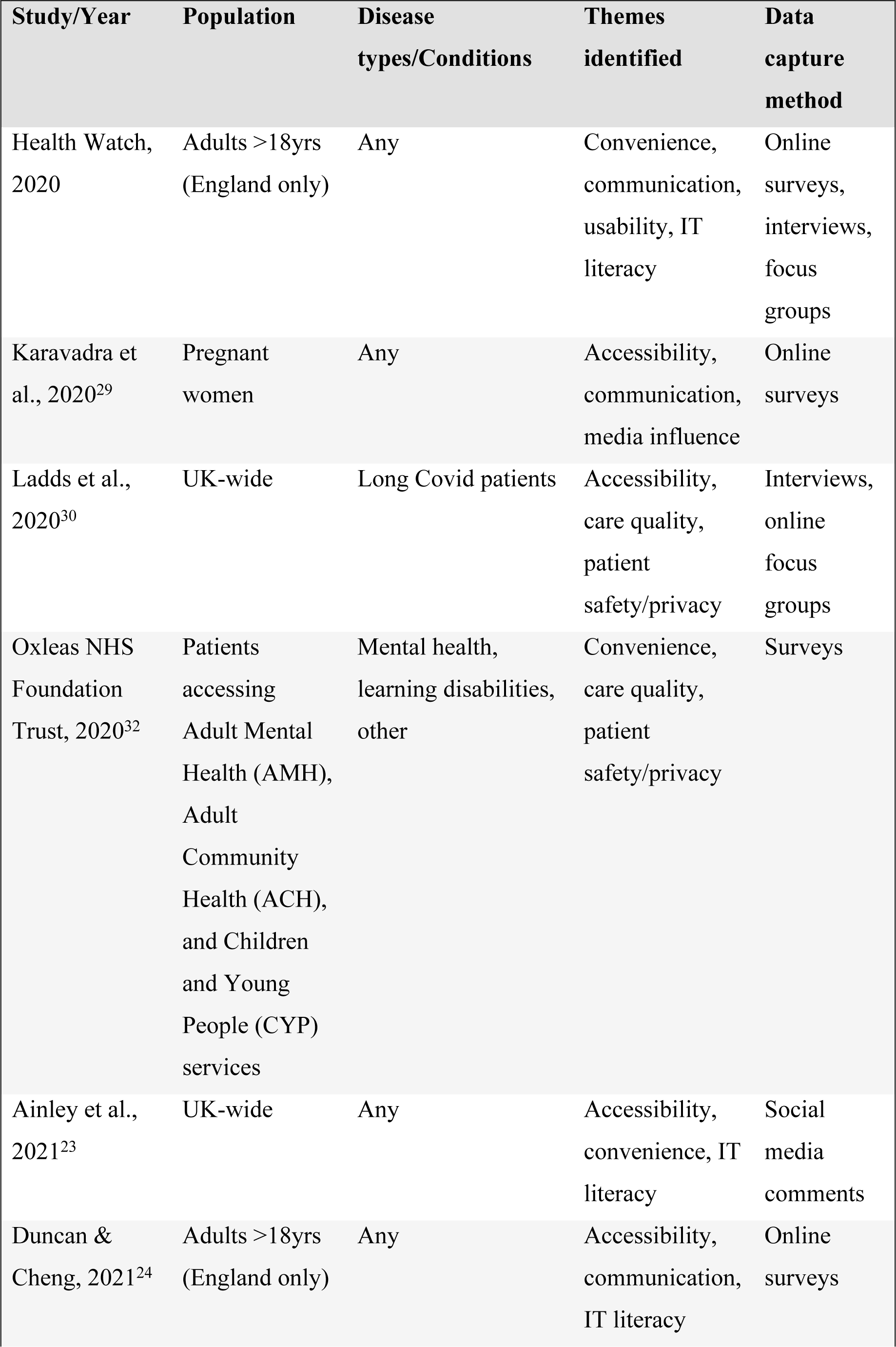
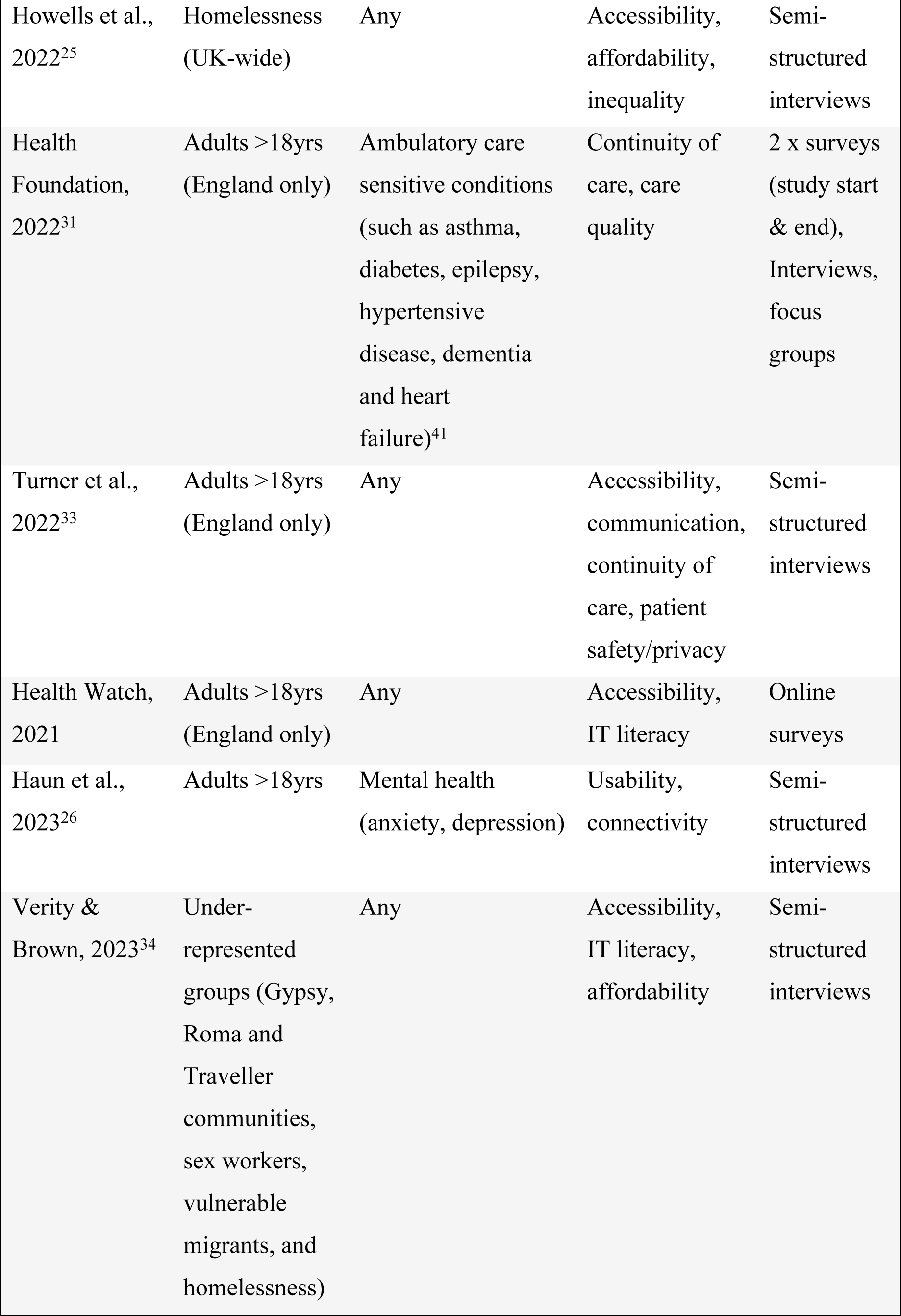
Study characteristics from earliest to most recently published.

**Table 4.**
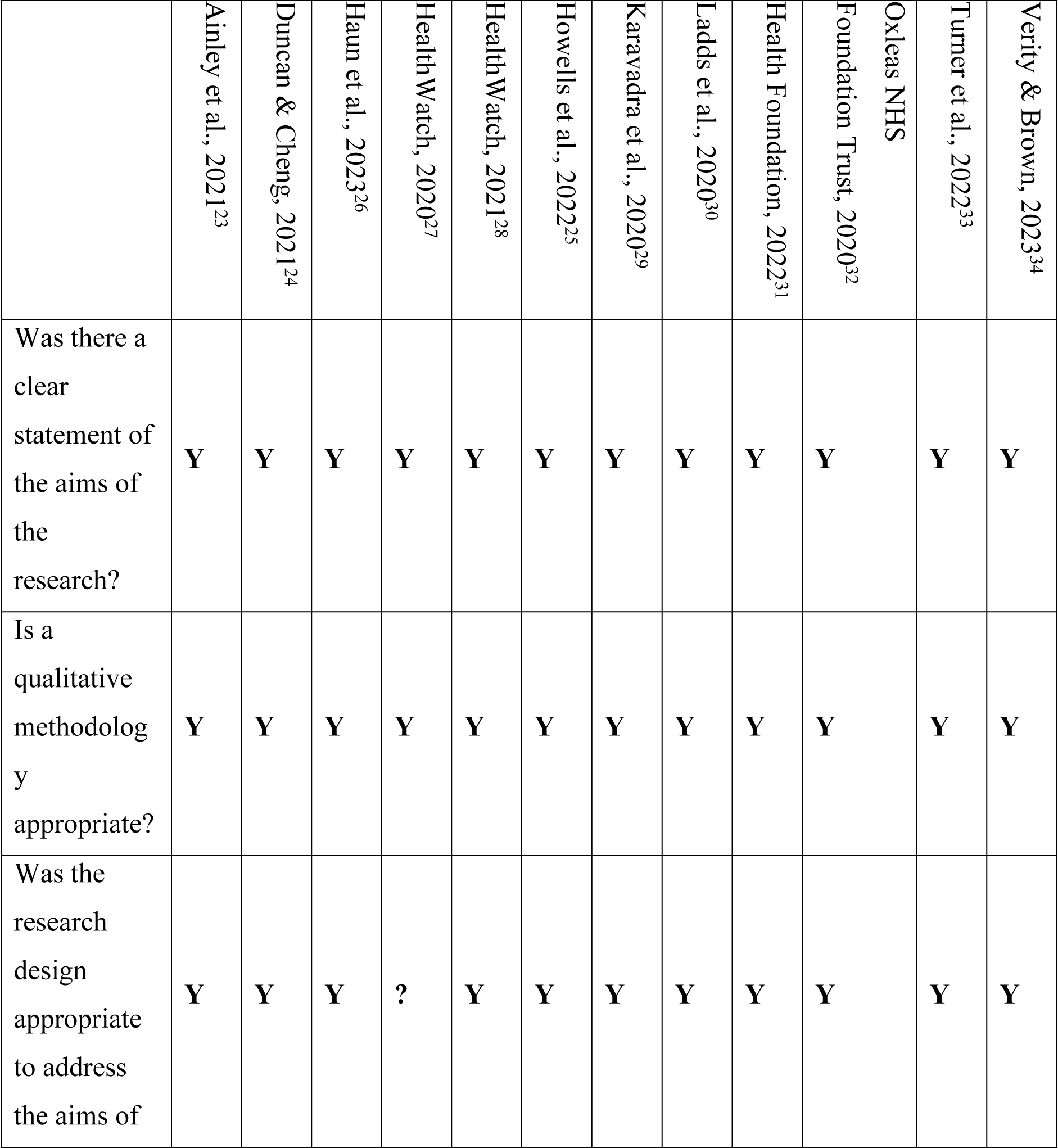
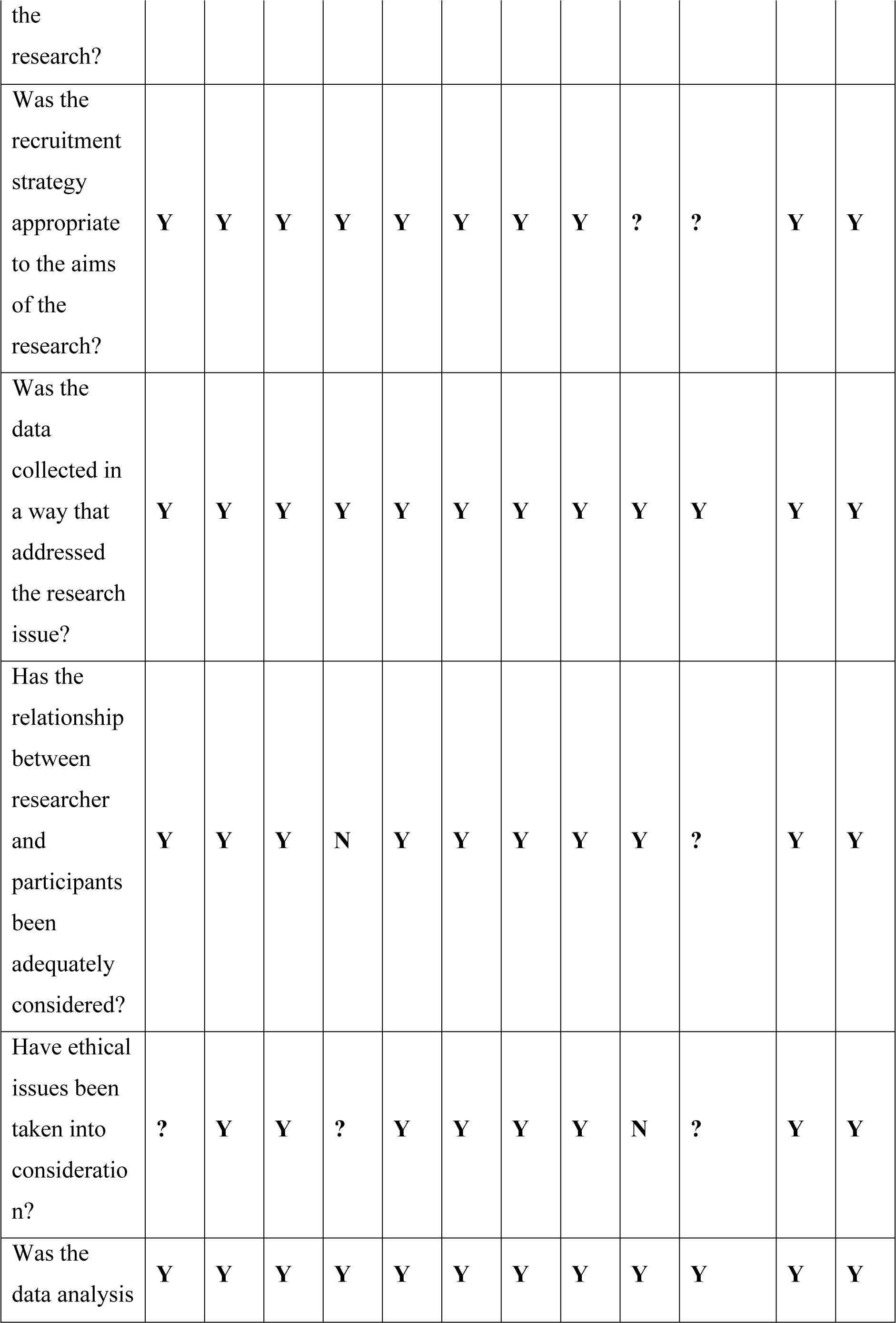
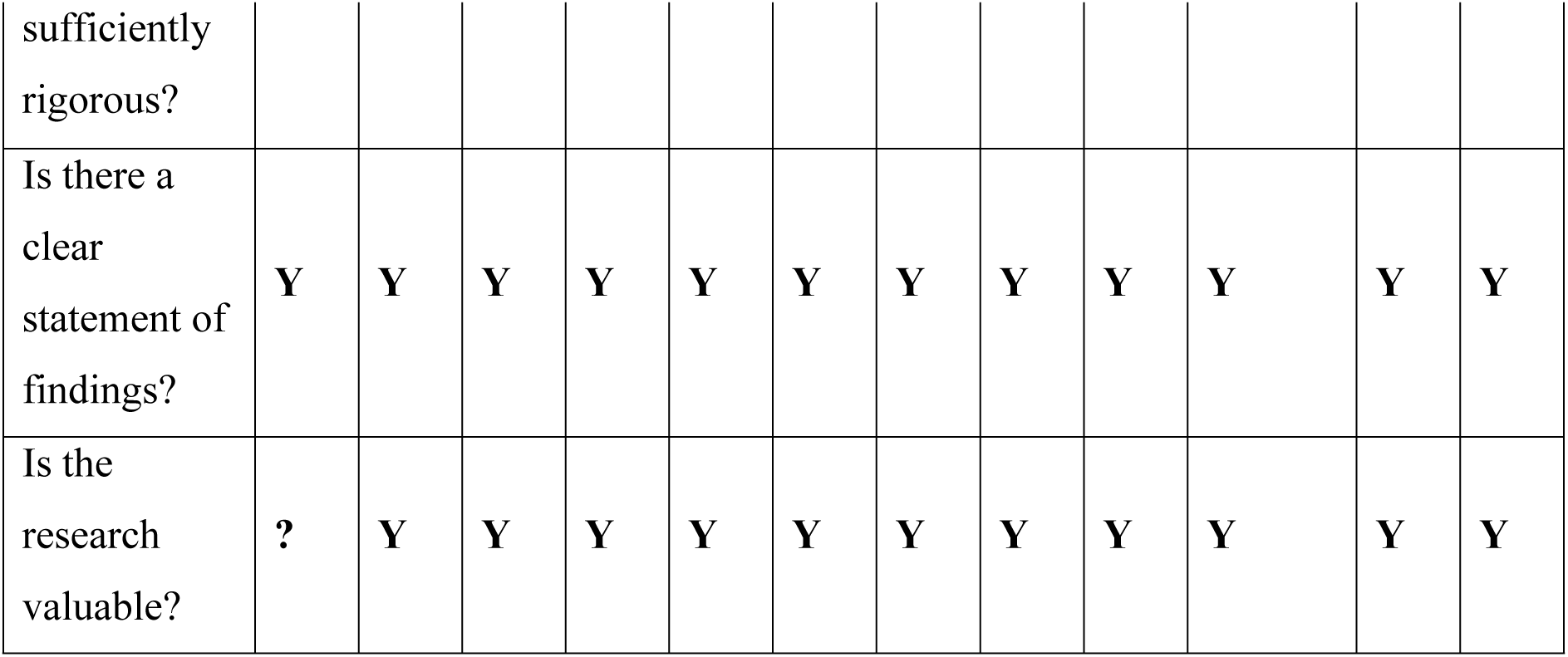
Summary of CASP quality appraisal of selected literature.

### Selection process

Results of the literature search were reviewed using the Critical Appraisal Skills Programme’s (CASP) checklist^16,17^.

## Data collection process

Qualitative data from selected literature were captured in QDA Miner^18^ to code themes from ingested data^19^. Data from literature sources came from the ‘results’ section of each publication and for grey literature were gathered from a complete review of the document. ‘Discussion’ sections were examined to indicate main themes. In papers with a mixed-methods approach patient experience data was carefully evaluated to ensure comments were not from quantitative aggregates of patient surveys that used Likert scaling for instance. Risk of bias was minimised using the CASP framework.

## Data Availability

Data derived from public domain resources

## References

1. Bakhai M, Atherton H. How to conduct written online consultations with patients in primary care. bmj. 2021 Feb 24;372.

2. Nijland N, van Gemert-Pijnen J, Boer H, Steehouder M, Seydel E. Evaluation of internet-based technology for supporting self-care: problems encountered by patients and caregivers when using self-care applications. Journal of medical Internet research. 2008 May 15;10(2):e957.

3. Baines R, Tredinnick-Rowe J, Jones R, Chatterjee A. Barriers and enablers in implementing electronic consultations in primary care: scoping review. Journal of Medical Internet Research. 2020 Nov 12;22(11):e19375.

4. England NH, Improvement NH. Advice on how to establish a remote ‘total triage’ model in general practice using online consultations. London: NHS England. 2020.

5. Edwards HB, Marques E, Hollingworth W, Horwood J, Farr M, Bernard E, Salisbury C, Northstone K. Use of a primary care online consultation system, by whom, when and why: evaluation of a pilot observational study in 36 general practices in South West England. BMJ open. 2017 Nov 1;7(11):e016901.

6. Eccles A, Hopper M, Turk A, Atherton H. Patient use of an online triage platform: a mixed-methods retrospective exploration in UK primary care. British Journal of General Practice. 2019 May 1;69(682):e336–44.

7. Miles DK, Stedman M, Heald AH. “Stay at Home, Protect the National Health Service, Save Lives”: A cost benefit analysis of the lockdown in the United Kingdom. International Journal of Clinical Practice. 2021 Mar;75(3):e13674.

8. Royal College of General Practitioners [internet]. GP consultations post-COVID should be a combination of remote and face to face, depending on patient need, says College. [cited 2023 Apr 11]. Available from: https://www.rcgp.org.uk/news/2021/may/gp-consultations-post-covid

9. Murphy M, Scott LJ, Salisbury C, Turner A, Scott A, Denholm R, Lewis R, Iyer G, Macleod J, Horwood J. Implementation of remote consulting in UK primary care following the COVID-19 pandemic: a mixed-methods longitudinal study. British Journal of General Practice. 2021 Mar 1;71(704):e166–77.

10. Page MJ, McKenzie JE, Bossuyt PM, Boutron I, Hoffmann TC, Mulrow CD, Shamseer L, Tetzlaff JM, Akl EA, Brennan SE, Chou R. The PRISMA 2020 statement: an updated guideline for reporting systematic reviews. International journal of surgery. 2021 Apr 1;88:105906.

11. Tugwell P, Tovey D. PRISMA 2020. Journal of Clinical Epidemiology. 2021 Jun 1;134:A5–6.

12. Page MJ, Moher D, McKenzie JE. Introduction to PRISMA 2020 and implications for research synthesis methodologists. Research synthesis methods. 2022 Mar;13(2):156–63.

13. Booth A, Cantrell A, Preston L, Chambers D, Goyder E. What is the evidence for the effectiveness, appropriateness and feasibility of group clinics for patients with chronic conditions? A systematic review.

14. Trinoskey J, Brahmi FA, Gall C. Zotero: A product review. Journal of Electronic Resources in Medical Libraries. 2009 Sep 9;6(3):224–9.

15. Erz H. Zotero 6: A Review.

16. Casp C. CASP qualitative checklist. Critical Appraisal Skills Programme. 2018 Apr 21.

17. Long HA, French DP, Brooks JM. Optimising the value of the critical appraisal skills programme (CASP) tool for quality appraisal in qualitative evidence synthesis. Research Methods in Medicine & Health Sciences. 2020 Sep;1(1):31–42.

18. Provalis Research [internet]. QDA Miner. [cited 2023 Mar 20]. Available from: https://provalisresearch.com/products/qualitative-data-analysis-software/freeware/

19. Adu P. A step-by-step guide to qualitative data coding. Routledge; 2019 Apr 5.

20. PRISMA [internet]. PRISMA Checklist. [cited 2023 Mar 25]. Available from: http://prisma-statement.org/PRISMAstatement/checklist.aspx?AspxAutoDetectCookieSupport=1

21. Carter P, Martin G. Challenges facing Healthwatch, a new consumer champion in England. International journal of health policy and management. 2016 Apr;5(4):259.

22. Department of Health and Social Care [internet]. Data saves lives: reshaping health and social care with data. [cited 2023 10 Apr]. Available from: https://www.gov.uk/government/publications/data-saves-lives-reshaping-health-and-social-care-with-data/data-saves-lives-reshaping-health-and-social-care-with-data

23. Ainley E, Witwicki C, Tallett A, Graham C. Using twitter comments to understand people’s experiences of UK health care during the COVID-19 pandemic: Thematic and sentiment analysis. Journal of Medical Internet Research. 2021 Oct 25;23(10):e31101.

24. Duncan LJ, Cheng KF. Public perception of NHS general practice during the first six months of the COVID-19 pandemic in England. F1000Research. 2021;10.

25. Howells K, Amp M, Burrows M, Brown J, Brennan R, Dickinson J, Jackson S, Yeung WL, Ashcroft D, Campbell S, Blakeman T. Remote primary care during the COVID- 19 pandemic for people experiencing homelessness: a qualitative study. British Journal of General Practice. 2022 Jul 1;72(720):e492–500.

26. Haun MW, Oeljeklaus L, Hoffmann M, Tönnies J, Wensing M, Szecsenyi J, Peters-Klimm F, Krisam R, Kronsteiner D, Hartmann M, Friederich HC. Primary care patients’ experiences of video consultations for depression and anxiety: a qualitative interview study embedded in a randomized feasibility trial. BMC Health Services Research. 2023 Dec;23(1):1–0.

27. Healthwatch.co.uk [internet]. The Doctor Will Zoom You Now: getting the most out of the virtual health and care experience. [cited 2023 Apr 12]. Available from: https://www.healthwatch.co.uk/sites/healthwatch.co.uk/files/The_Dr_Will_Zoom_You_Now_-_Insights_Report.pdf

28. Healthwatch.co.uk [internet]. GP access during COVID-19. [cited 2023 Apr 15]. Available from: https://www.healthwatch.co.uk/sites/healthwatch.co.uk/files/20210215%20GP%20access%20during%20COVID19%20report%20final_0.pdf

29. Karavadra B, Stockl A, Prosser-Snelling E, Simpson P, Morris E. Women’s perceptions of COVID-19 and their healthcare experiences: a qualitative thematic analysis of a national survey of pregnant women in the United Kingdom. BMC pregnancy and childbirth. 2020 Dec;20(1):1–8.

30. Ladds E, Rushforth A, Wieringa S, Taylor S, Rayner C, Husain L, Greenhalgh T. Persistent symptoms after Covid-19: qualitative study of 114 “long Covid” patients and draft quality principles for services. BMC health services research. 2020 Dec;20(1):1–3.

31. The Health Foundation [Internet]. Increasing continuity of care in general practice programme. [cited 2023 Apr 12]. Available from: https://www.health.org.uk/sites/default/files/2022-12/continuity_of_care_final_independent_evaluation_mixedmethodsevalreport_2022.pdf

32. Oxleas NHS Foundation Trust [internet]. Patient Experience of Remote Consultations during the COVID-19 Pandemic. [cited 2023 Apr 15]. Available from: https://healthinnovationnetwork.com/wp-content/uploads/2022/01/Remote-experience-reportV7.pdf

33. Turner A, Morris R, Rakhra D, Stevenson F, McDonagh L, Hamilton F, Atherton H, Farr M, Blake S, Banks J, Lasseter G. Unintended consequences of online consultations: a qualitative study in UK primary care. British Journal of General Practice. 2022 Feb 1;72(715):e128–37.

34. Verity A, Brown VT. Inclusion health patient perspectives on remote access to general practice: a qualitative study. BJGP open. 2023 Apr 19.

35. Braun V, Clarke V. Using thematic analysis in psychology. Qualitative research in psychology. 2006 Jan 1;3(2):77–101.

36. Byrne D. A worked example of Braun and Clarke’s approach to reflexive thematic analysis. Quality & quantity. 2022 Jun;56(3):1391–412.

37. Anderson M, Pitchforth E, Asaria M, Brayne C, Casadei B, Charlesworth A, Coulter A, Franklin BD, Donaldson C, Drummond M, Dunnell K. LSE–Lancet Commission on the future of the NHS: re-laying the foundations for an equitable and efficient health and care service after COVID-19. The Lancet. 2021 May 22;397(10288):1915–78.

38. The Kings Fund [internet]. Public satisfaction with the NHS and social care in 2022: Results from the British Social Attitudes survey. [cited 2023 Apr 14]. Available from: https://www.kingsfund.org.uk/publications/public-satisfaction-nhs-and-social-care-2022

39. Gille F, Smith S, Mays N. Why public trust in health care systems matters and deserves greater research attention. Journal of health services research & policy. 2015 Jan;20(1):62–4.

40. Goldenberg MJ. Vaccine hesitancy: public trust, expertise, and the war on science. University of Pittsburgh Press; 2021 Mar 30.

41. NHS England [internet]. Emergency admissions for Ambulatory Care Sensitive Conditions – characteristics and trends at national level. [cited 2023 Apr 15]. Available from: https://www.england.nhs.uk/wp-content/uploads/2014/03/red-acsc-em-admissions-2.pdf

42. NHS Health Research Authority [internet]. Public involvement in a pandemic: Lessons from the UK COVID-19 public involvement matching service. [cited 2023 Apr 14]. Available from: https://s3.eu-west-2.amazonaws.com/www.hra.nhs.uk/media/documents/8948_Public_Involvement_in_Pandemic_Research_Report_V9_-_Accessible.pdf

